# Distinct Synaptic Excitation–Inhibition Mechanisms Underlie Clinically Defined Seizure Onset Patterns

**DOI:** 10.64898/2026.03.25.26349297

**Authors:** Isa Dallmer-Zerbe, Anna Pidnebesna, Jaroslav Hlinka

## Abstract

Epileptic seizures exhibit marked phenotypic heterogeneity that reflects distinct underlying network mechanisms, yet these differences are incompletely captured by current clinical classifications. Computational models offer a principled approach to infer latent excitation–inhibition dynamics from intracranial EEG, enabling mechanism-informed seizure characterization. We analyzed 205 seizures from 15 patients with drug-resistant epilepsy from the European Epilepsy Database, covering seven clinically annotated seizure onset patterns. Using the Wendling neural mass model, we fitted five-second iEEG segments by optimizing synaptic excitation and inhibition parameters across four temporal windows spanning 60 s before to 25 s after seizure onset. Model-derived excitation–inhibition changes distinguished seizure types significantly above chance. Classification performance was strongest when combining excitation and inhibition parameters, with peridendritic inhibition being the single most discriminative parameter. Seizure-type–specific signatures were detectable not only during seizure onset and within seizure onset zones, but already during interictal periods and in non-onset channels, indicating that seizure mechanisms are preconfigured tens of seconds before clinical onset and extend beyond focal onset regions. Although all seizure types showed increases in both excitation and inhibition during seizure transition, their timing and magnitude differed systematically. In particular, our study supports and extends prior evidence that high-amplitude slow (HAS) seizures are driven by localized hyperexcitation within the seizure onset zone, whereas low-amplitude fast (LAF) seizures arise from inhibition-driven network mechanisms. Excitation–inhibition signatures were further linked to individual patient characteristics and surgical outcomes, highlighting their potential clinical relevance.

## INTRODUCTION

The morphology of focal seizure onsets in intracranial EEG (iEEG) recordings of epilepsy patients exhibits large variability, including high-amplitude/low-frequency periodic spikes, low-voltage fast activity, spike-and-wave discharges, and sharp transients below 13 Hz (Abdallah et al., 2024; Perucca et al., 2014). These patterns reflect underlying electrophysiological mechanisms and can be classified via signal feature extraction and computational modeling (Perucca et al., 2014; Wang et al., 2017).

The recognized determinants of seizure onset patterns are diverse, including both the underlying pathology and the spatial organization of the epileptogenic zone (Lagarde et al., 2019). For example, low-frequency high-amplitude periodic spikes are specific to the mesial temporal lobe, which is associated with mesial temporal atrophy/sclerosis (Perucca et al., 2014). In focal cortical dysplasia and neurodevelopmental tumors, on the other hand, six seizure onset patterns have been described, with low-voltage fast activity being the most prevalent pattern (Lagarde et al., 2016). It has further been shown that certain onset patterns in focal refractory epilepsies correlate with surgical outcomes (Lagarde et al., 2016). For example, the presence of low-voltage fast activity in the seizure onset pattern has often been associated with a better postsurgical outcome (Lagarde et al., 2016; Wetjen et al., 2010). Furthermore, seizure onset patterns have been found to differ significantly in terms of the extent of the seizure onset zone (SOZ), with low-voltage fast activity associated with a larger SOZ (Perucca et al., 2014). Interestingly, seizure onset patterns are not limited to the seizure onset zone but occur also in other brain regions. For example, regions of seizure spread have been linked with sharp activity at ≤13 Hz and low-voltage fast activity (Perucca et al., 2014).

Given the differences in underlying pathology, spatial extent and associated surgery outcome, different types of seizure onsets might imply different types of necessary treatment parameters. Therefore, seizure onset patterns are being extensively studied to understand their role in defining the epileptogenic zone and predicting surgical outcomes.

As for the two most commonly observed onset types: the hypersynchronous onset (low-frequency high-amplitude periodic spikes at a frequency <2 Hz) and the low-voltage fast activity onset (mainly in beta and gamma range) (Chang et al., 2024), clinical and experimental studies have identified distinct cellular and network mechanisms. *In vivo* and *in vitro* studies indicate that hypersynchronous seizure onset is driven by overexcitation and a breakdown of surround inhibition in SOZ, enabling initially localized hyperactivity in principal neurons to propagate (see Devinsky et al., 2018, Fig. 4; Koehling et al., 2016). In contrast, the surrounding cortical tissue in low-voltage fast activity onset is characterized by comparatively low excitability, which may limit rapid seizure spread (Schevon et al., 2012; Wang et al., 2017). Instead, seizures with a low-voltage onset emerge through network activation predominantly driven by inhibitory mechanisms, without the immediate engagement of *principal neurons* (see Devinsky et al., 2018, Fig. 4; de Curtis & Avoli, 2016; Grasse et al., 2013). Consistently, cortical excitability, measured via cortico-cortical evoked potentials, is reduced in the SOZ of seizures with low-voltage onset compared to those with hypersynchronous onset (Enatsu et al., 2012). Adding further complexity, ictogenesis is also modulated by non-synaptic and non-neuronal factors (Ayala et al., 1970).

Computational modeling has significantly advanced the study of seizure onset patterns in epilepsy by providing frameworks to simulate and analyze neuronal dynamics underlying seizure initiation. Established models reproducing epileptic activity focused on single neuron and neuronal population excitability and synaptic interactions, highlighting the role of ionic currents, network connectivity and excitation/inhibition balance in seizure generation (Jirsa et al., 2014; Wendling et al., 2002). Recent studies have expanded to large-scale brain networks that incorporate patient-specific structural and functional data (see Dallmer-Zerbe et al. 2023 for a review). Moreover, computational modeling has offered new ways to classify seizure onset types, based on their associated on- and offset dynamics (Saggio et al., 2020). As for the low-voltage fast activity onset and the hypersynchronous onset types, a recent study by Wang et al. (2017) distinguished them by differing tissue excitability and spatial organization. They find that in hypersynchronous onset type seizures (here “HAS: High Amplitude Slow”) a localized trigger induces abrupt, widespread seizure recruitment across cortical tissue that exhibits elevated baseline excitability, while low-voltage fast activity onset (here “LAF: Low Amplitude Fast” seizures occur through multiple separate regions of abnormal activity that progressively merge and expand into adjacent cortical areas characterized by normal baseline excitability levels (Wang et al., 2017; see also Shokooh et al. 2023,). Wendling et al. (2002, 2005), as well as others, further highlight the role of inhibitory transmission as a key contributor to LAF type seizure generation.

Despite recent advances, challenges remain in capturing the full heterogeneity of seizure dynamics and understanding the involved brain mechanisms in seizure initiation across patients and clinical conditions, underscoring the need for integrative modeling frameworks informed by personalized parameters (Goodfellow et al., 2016).

This study used signal feature analysis and computational modeling to analyze intracranial EEG recordings around seizure onset. Building on the previous work of Wendling et al. (2002, 2005) and Dallmer-Zerbe et al. (2023) the goal was to demonstrate that different seizure onset patterns are linked with distinct profiles of changes in synaptic communication related to excitation/inhibition balance, supported by personalized computational modeling. We further: 1) show that seizure onset patterns are not limited to the time of seizure onset and clinically marked SOZ, 2) explore the mechanisms of LAF and the HAS type seizure onsets in more detail, and 3) consider patient-specific variables such as epilepsy localization and surgery outcome.

## METHODS

### Dataset

The dataset consisted of 15 intracranially recorded patients from the European Epilepsy database (https://www.epilepsy-database.eu/), developed within the EU-funded EPILEPSIAE project conducted at three epilepsy centers in Portugal (Coimbra), France (Paris), and Germany (Freiburg). It provides (almost) continuous iEEG recordings, with varying numbers of recorded seizures across patients, sampled at rates between 250 and 1024 Hz. It further includes rich clinical metadata, such as standardized annotations for seizure occurrences, types of observed seizure onset pattern per seizure used in this study and entails comprehensive patient information covering aspects such as medical history, seizure semiology, administered antiepileptic drugs, and outcome and description of surgical interventions. For the iEEG recording grid, strip and depth electrodes were used. Further dataset information can be found in (Klatt et al., 2012). The recordings used in this study were invasive EEG recordings from the University Hospital Freiburg. Patient characteristics are listed in Table 1.

**Table 1.**
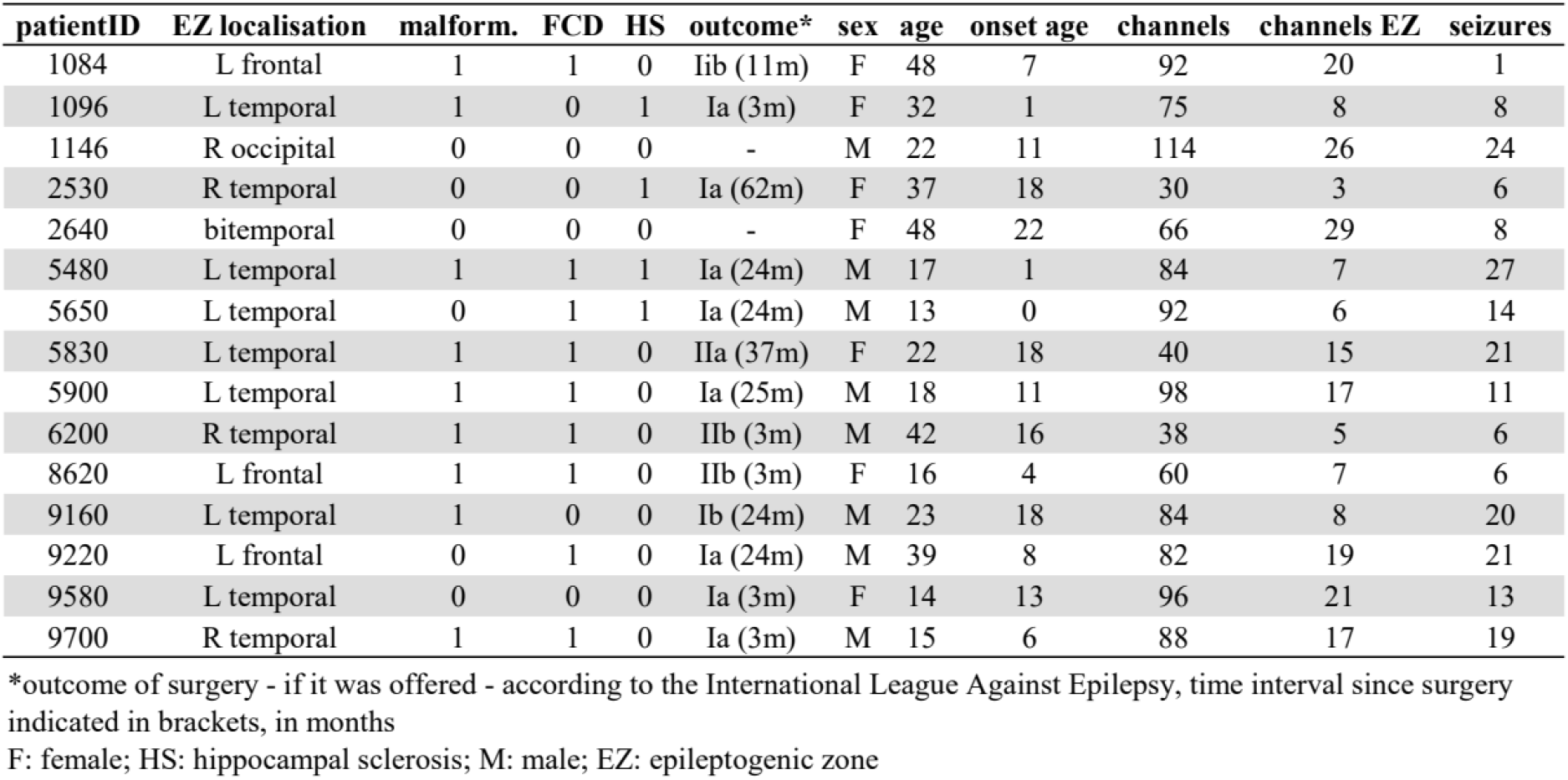
Individual patient characteristics of the EuEpi dataset.

| patientID | EZ localisation | malform. | FCD | HS | outcome* | sex | age | onset age | channels | channels EZ | seizures |
| --- | --- | --- | --- | --- | --- | --- | --- | --- | --- | --- | --- |
| 1084 | L frontal | 1 | 1 | 0 | Iib (11m) | F | 48 | 7 | 92 | 20 | 1 |
| 1096 | L temporal | 1 | 0 | 1 | Ia (3m) | F | 32 | 1 | 75 | 8 | 8 |
| 1146 | R occipital | 0 | 0 | 0 | - | M | 22 | 11 | 114 | 26 | 24 |
| 2530 | R temporal | 0 | 0 | 1 | Ia (62m) | F | 37 | 18 | 30 | 3 | 6 |
| 2640 | bitemporal | 0 | 0 | 0 | - | F | 48 | 22 | 66 | 29 | 8 |
| 5480 | L temporal | 1 | 1 | 1 | Ia (24m) | M | 17 | 1 | 84 | 7 | 27 |
| 5650 | L temporal | 0 | 1 | 1 | Ia (24m) | M | 13 | 0 | 92 | 6 | 14 |
| 5830 | L temporal | 1 | 1 | 0 | IIa (37m) | F | 22 | 18 | 40 | 15 | 21 |
| 5900 | L temporal | 1 | 1 | 0 | Ia (25m) | M | 18 | 11 | 98 | 17 | 11 |
| 6200 | R temporal | 1 | 1 | 0 | IIb (3m) | M | 42 | 16 | 38 | 5 | 6 |
| 8620 | L frontal | 1 | 1 | 0 | IIb (3m) | F | 16 | 4 | 60 | 7 | 6 |
| 9160 | L temporal | 1 | 0 | 0 | Ib (24m) | M | 23 | 18 | 84 | 8 | 20 |
| 9220 | L frontal | 0 | 1 | 0 | Ia (24m) | M | 39 | 8 | 82 | 19 | 21 |
| 9580 | L temporal | 0 | 0 | 0 | Ia (3m) | F | 14 | 13 | 96 | 21 | 13 |
| 9700 | R temporal | 1 | 1 | 0 | Ia (3m) | M | 15 | 6 | 88 | 17 | 19 |

\*outcome of surgery - if it was offered - according to the International League Against Epilepsy, time interval since surgery indicated in brackets, in months
F: female; HS: hippocampal sclerosis; M: male; EZ: epileptogenic zone

For each marked seizure in the dataset, we extracted, if available, the time interval of 60s before until 25 seconds after clinically marked seizure onset, leaving us with a total of n = 205 seizures. We centered the data of each extracted seizure recording around zero by deducting the mean across all data points per recording channel, to remove DC-related differences between recordings. We then dissected the channel-specific seizure recordings into five-second segments. Finally, we assigned each segment a type label based on its timing with regards to clinically marked seizure onset; “interictal”: 60s to 30s before seizure onset, “preonset”: 30s to 0s before seizure onset, “onset”: 0s to 10s after seizure onset; “ictal”: 10s to 25s after seizure onset, leading to a total of 30 / 5 = 6 interictal segments, 6 preonset, 2 onset, and 3 ictal segments, per seizure recording. Note, that each seizure was marked by clinicians regarding the observed seizure onset pattern, namely “a: rhythmic alpha waves” (n = 4), “b: rhythmic beta waves” (n = 57), “l: low amplitude fast activity” (n = 70), “p: polyspikes” (n = 18), “r: repetitive spiking” (n = 41), “s: rhythmic sharp waves” (n = 12), “t: rhythmic theta waves” (n = 3). Furthermore, seizures were clinically marked as being either “CP: complex partial” (n = 61), “SG: secondarily generalized” (n = 23), “SP: simple partial” (n = 52), or “UC: unclassified” (n = 69), allowing further possible investigation in the future.

### Model simulations

We simulated the Wendling neural mass model (Wendling et al., 2002, 2005) starting from the implementation by (Fietkiewicz & Loparo, 2016), using the parameters listed in Table 2. The three main parameters of the model considered in this study correspond to the average excitatory synaptic gain (A), exerted by the pyramidal cell population, to the average slow inhibitory synaptic gain (B), commonly attributed to the Somatostatin-expressing (SOM+) interneurons, and to the average fast inhibitory synaptic gain (G), attributed to the Parvalbumin-expressing (PV+) interneurons (Wendling et al., 2005). Upon changing these parameters, the model can reproduce different types of epileptic activity (Wendling et al., 2002, 2005, Dallmer-Zerbe et al. 2023). We fit these parameters to individual data segments of each recording channel per seizure to get an estimate of A, B and G variations in time. For the fitting procedure, we generated n = 141659 five-second simulated time series, representing the EEG recording obtained for the 141659 unique sets of A, B, and G values. The parameters were systematically varied within their respective biological boundaries (A: [2 14], B: [1 25], G: [1 30]), with a resolution of 0.25 for parameter A and 0.5 for parameter B and G.

**Table 2.**
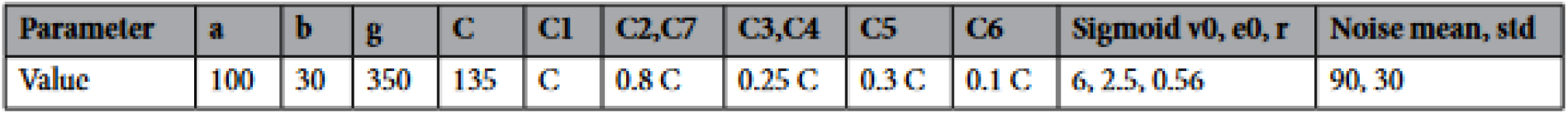
Model parameters used for simulations.

### Model fitting procedure

From all the simulated segments with different A, B, and G, we identified the one segment that most resembles the current real data segment, assigning each real data segment the one (A, B, G) configuration which led to the most similar model output. Best resemblance was assessed via minimizing the distance between the real segment’s feature values and the simulated segments’ feature values (11 signal features, including power band and spike detecting features, mean, variance, line length and autocorrelation, for details see *Feature calculation and normalization* in Dallmer-Zerbe et al. 2023). The fitting procedure followed the steps (1) feature calculation, (2) principal component analysis, and (3) A, B, G fitting. Steps (1) and (2) were the same as in the Dallmer-Zerbe et al. 2023, including the projection into the prototype-defined PCA space. However, in step (2) in this study, segment features were z-normalized using the feature-specific mean and standard deviation of interictal segments’ features only (both model and real data segments), to best capture the seizure-related changes with regard to background/interictal iEEG. For the real data we used the interictal segments of the respective recording. For normalization of the model data we used 100 interictal segments, simulated with parameters set to A = 3.5, B = 13.2, and G = 10.76 (Wendling et al., 2005). As an estimate of current levels of excitation (A), slow inhibition (B) and fast inhibition (G) levels, respectively, for each of the real data segments, we used the (A, B, G) configuration which led to the most similar model output in step (3) (as assessed via feature comparison) (Fig. 1).

**Figure 1:**
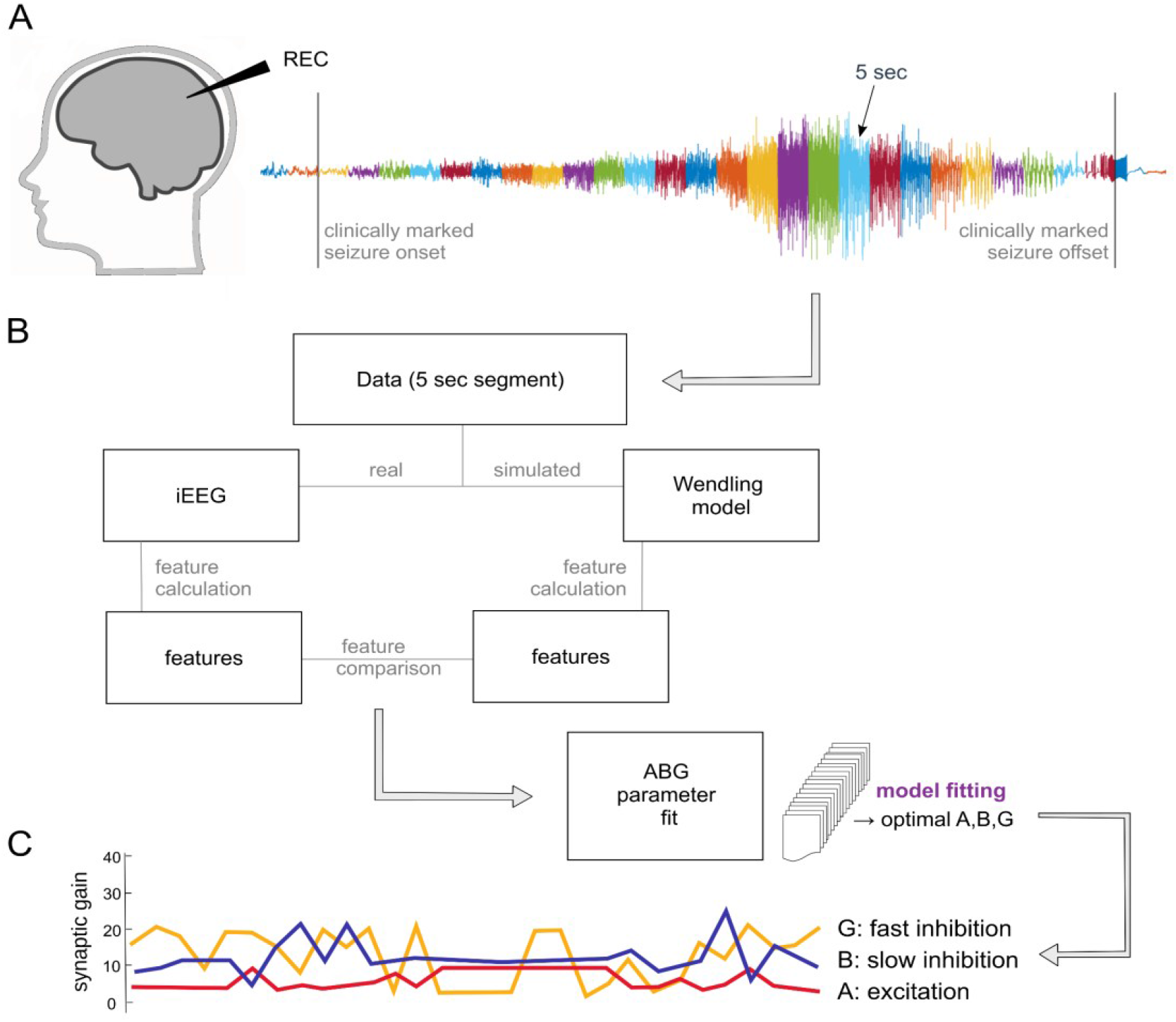
Data analysis and model fitting procedure. **(A)** Example single-channel seizure recording of patient 5900 with color-coded 5-second data segmentation. **(B)** Scheme of the data analysis and model fitting procedure. **(C)** Resulting predicted A, B, G evolution for the example recording in A.

### Description of the data analyses

A, B, and G values were averaged over segments for each type (interictal, preonset, onset, and ictal), and then across channels per seizure. In a first analysis, we further averaged per patient, to explore the general synaptic mechanisms with regards to changes in excitation and inhibition during the interictal, preonset, onset and ictal intervals. An increasing trend in A, B, and G parameters was statistically assessed via one-sided Wilcoxon rank sum testing of each parameter for the interictal vs ictal interval. *p*-values were FDR corrected to account for multiple comparisons (Benjamini-Yekutieli procedure, 3 comparisons).

In a second analysis, we were interested to see whether the clinically marked seizure onset pattern per seizure could be predicted above chance level based on the seizure-specific A, B, G evolution (assuming that there could be different mechanisms guiding the transition to seizures for different types of seizure onset patterns). To this end, we used a linear support vector classifier *SVC (kernel = ‘linear’)* from the *scikit-learn* library (version *1*.*3*.*0*) in Python (version *3*.*12*.*1*), with the *class_weight* parameter set to ‘balanced’ to account for class imbalance in the dataset (different amounts of seizures per type). Before the classification, all the features were z-transformed. To assess the classification performance, we conducted leave-one-out cross-validation at the seizure level: every individual seizure was left out in turn, while all the others were used for model training; then, the model was used to predict the label of the left-out seizure. We would like to point out that leave-one-patient-out cross-validation strategy would be preferable; however, due to the high number of seizure types and strong class imbalance, we did not have enough data to rely on the obtained estimates. Nevertheless, the results for leave-one-patient-out cross-validation are presented in the Supplementary Material, see S3 and S4. Classification performance was measured using balanced accuracy (which is the average of recall obtained on each class). We further calculated p-values to determine whether the achieved balanced accuracies were significant. We computed p-values by running the classification on randomly permuted labels (500 permutations) and calculating the proportion of simulations with a higher accuracy than the one for the real data.

First, to test the general mechanistic differences of clinically marked seizure onset patterns, we applied the above-described classification procedure to the seizure-specific A, B, G evolutions (“Trajectory”: including all 4 time points), averaged across All channels per seizure. Then, in a set of post-hoc analyses, we tested which of the time points (interictal, preonset, onset, ictal), brain locations (SOZ, Other, All) and excitation/inhibition parameters (A, B, G) were most critical to the differentiation of the seizure types. For this, we compared the classification performance when limiting the analysis to single time points, brain locations of parameters. In particular, we investigated the time point of seizure onset in SOZ, as we believed the classification should perform best there, and the interictal time point in Other as a comparison, where we expected the lowest classification performance. Furthermore, we studied the classification performance for A, B, and G parameters separately (Supplement S2). We thereby aimed to answer the question, which synaptic population (Pyramidal cells: excitatory synaptic gain A, Parvalbumin-expressing (PV) interneurons: fast inhibitory synaptic gain G, and Somatostatin-expressing (SST) slow inhibitory synaptic gain B) would be most discriminative between the different types of seizure transitions. The resulting *p*-values were FDR corrected on the alpha = 0.05 level via Benjamini–Hochberg procedure for each parameter-specific results table, i.e. separately for (ABG), A, B and G. All the results tables were of a shape of 5 time points (interictal, preonset, onset, ictal and full trajectory) x 3 brain location (All, Other, SOZ).

To exemplify the mechanistic insight gained by the model fitting procedure, we then compared the onset mechanisms of two most common seizure onset types: “l: low amplitude fast activity” (corresponding to low-voltage or LAF onset in the introduction) and “r: repetitive spiking” (corresponding to hypersynchronous or HAS onset in the introduction) in more detail. Based on the literature (see introduction) we tested the following directed hypotheses: 1) higher excitation (A) during seizure onset in SOZ of “r” type seizures, reflecting hyperexcitability mechanisms overcoming surround inhibition, as compared to “l” type seizures, where no immediate principal neuron engagement is expected. 2) Higher inhibition (B and G) at the onset of “l” type seizures, reflecting predominantly inhibition-driven mechanisms, as compared to “r” type seizures, where no inhibition changes or even inhibition failure is expected. As before, we used one-sided Wilcoxon rank sum testing and Benjamini-Yekutieli-FDR correction (3 comparisons: A_r_> A_l_, B_r_< B_l_, G_r_ < G_l_). We further tested SOZ - Other differences in parameter values averaged across all four-time windows separately for “r” and “l” types using one-sided Wilcoxon signed rank testing and FDR correction (3 × 2 comparisons, e.g. A_r_ (SOZ - Other) > 0, A_L_ (SOZ - Other) > 0, B_r_ (SOZ - Other) > 0, etc.). Based on the literature, we expected that excitation changes in “r” type seizures should be mostly localized in the SOZ, thus yielding A_r_ (SOZ - Other) > 0 significant, while inhibition changes in “l” type seizures would be network-wide, yielding B_l_/G_l_(SOZ - Other) > 0 non-significant.

Finally, we assessed a potential relationship of the identified differences between patients with individual patient characteristics (see Table 1). As the patient sample was too small and thus statistical power too low, this analysis was of a descriptive nature only and entailed no significance testing.

## RESULTS

We studied model-identified excitation/inhibition levels during seizure transition (assessed in four-time windows: interictal, preonset, onset, ictal; as defined with regards to clinically marked seizure onset time), in “All”, “SOZ”, or “Other”, recording channels per seizure. We expected a general increase in excitation/inhibition from interictal to ictal. Moreover, different seizure types were hypothesized to be characterized by distinct excitation–inhibition dynamics (A: excitation, B: slow inhibition, G: fast inhibition) during seizure transition, with the best discrimination expected for SOZ channels at the time of seizure onset (which is the time interval used by the clinical experts when visually discriminating seizure type).

Fig. 2A shows an exemplary data recording, Fig. 2B its resulting model-fit for the excitation parameter A, Fig. 2C the averaged A values per time interval across channels and finally, Fig. 2D, the grand-average evolution of the excitation/inhibition levels during seizure transition across the 15 patients in this study. Excitatory, slow and fast inhibitory synaptic gain parameters A, B, and G were each averaged across all channels and then across seizures per patient. We find that overall, both excitation and inhibition increased during seizure transitions (one-sided Wilcoxon signed rank test of (ictal-interictal) > 0 with *p*_*FDR*_ = .000 for A; B: *p*_*FDR*_ = .004; and G: *p*_*FDR*_ = .000).

**Figure 2:**
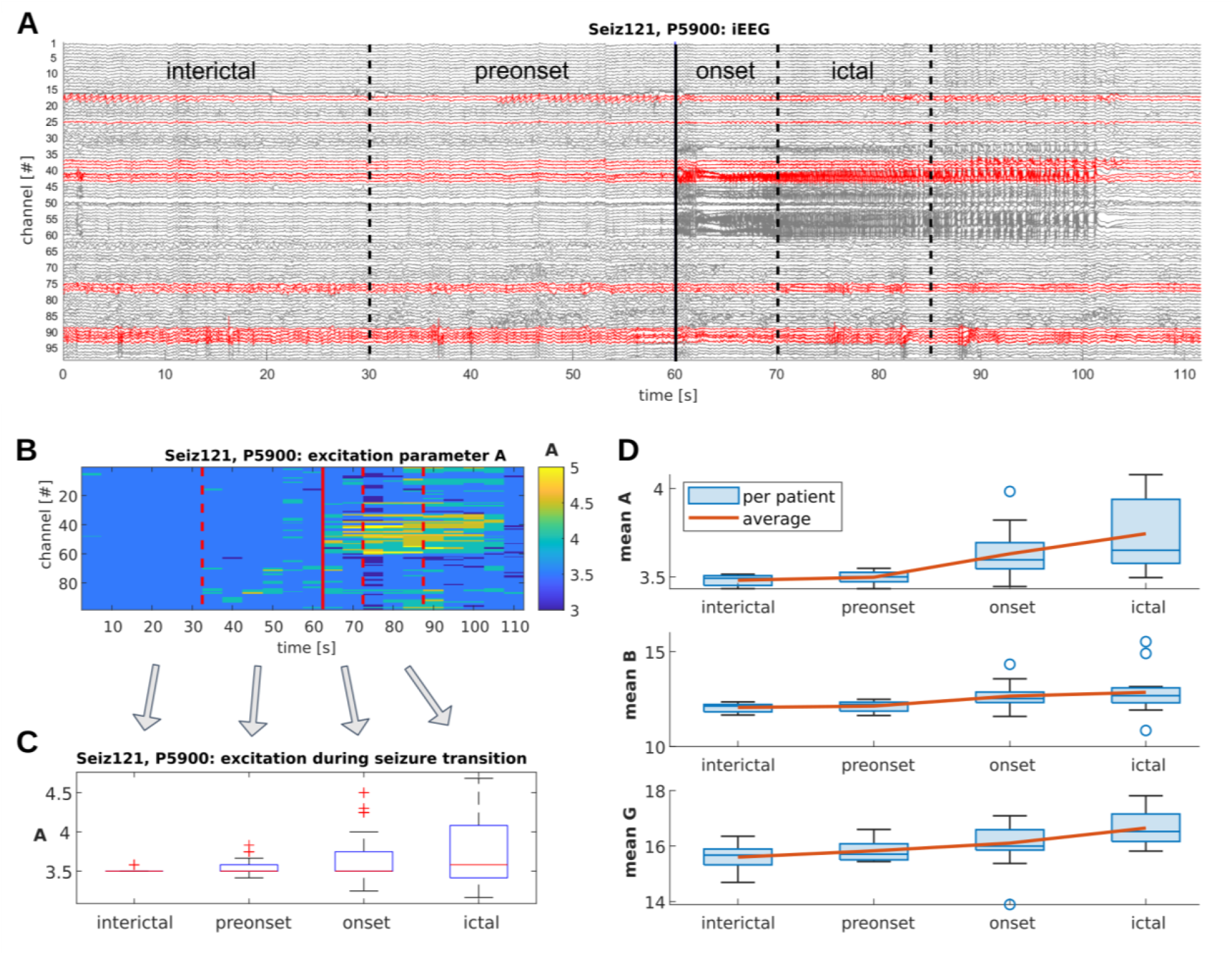
Excitation (A), slow inhibition (B), and fast inhibition (G) levels during seizure transition. **(A)** Example seizure recording of patient 5900. The solid and dashed lines mark the respective end of each interval: interictal, preonset, onset, ictal. The black solid line indicates clinically marked seizure onset, which always occurred one minute after the beginning of the selected data window. Dashed lines correspond to (arbitrarily) chosen segmentation between intervals. The different recording channels are colored with regards to their location either inside or outside of seizure onset zone (SOZ: red, Other: black), as determined by the clinical experts. **(B)** Model fitting result example for excitatory synaptic gain parameter A determined for every 5 second data segment of every recording channel for the seizure recording in panel A. The red lines mark the end of interictal, preonset, onset, and ictal intervals as in panel A. Note that the values of excitation parameter A are variable across channels and time. The seizure-related patterns in the raw signal in channels 40 to 60 (see panel A), overlap with high values in excitation in panel B. **(C)** The estimates were then averaged across four time windows (interictal, preonset, onset, ictal) per channel, to show the general evolution of excitation levels during seizure transition. **(D)** Average excitation (A), slow inhibition (B), and fast inhibition (G) change from interictal to ictal transition across patients (after averaging across channels and seizures per patient). We observe an overall increasing trend in excitation and fast inhibition levels.

In the next step, we studied the excitation/inhibition dynamics for different seizure types (Fig. 3). Here, instead of averaging across seizures per patient as in Fig. 1D, we averaged the identified A, B and G values across seizures per type (Fig. 3A and B). We then tested the statistical differences between grand average values across seizures for all seven types labeled in the dataset using a classifier (see Methods).

**Figure 3:**
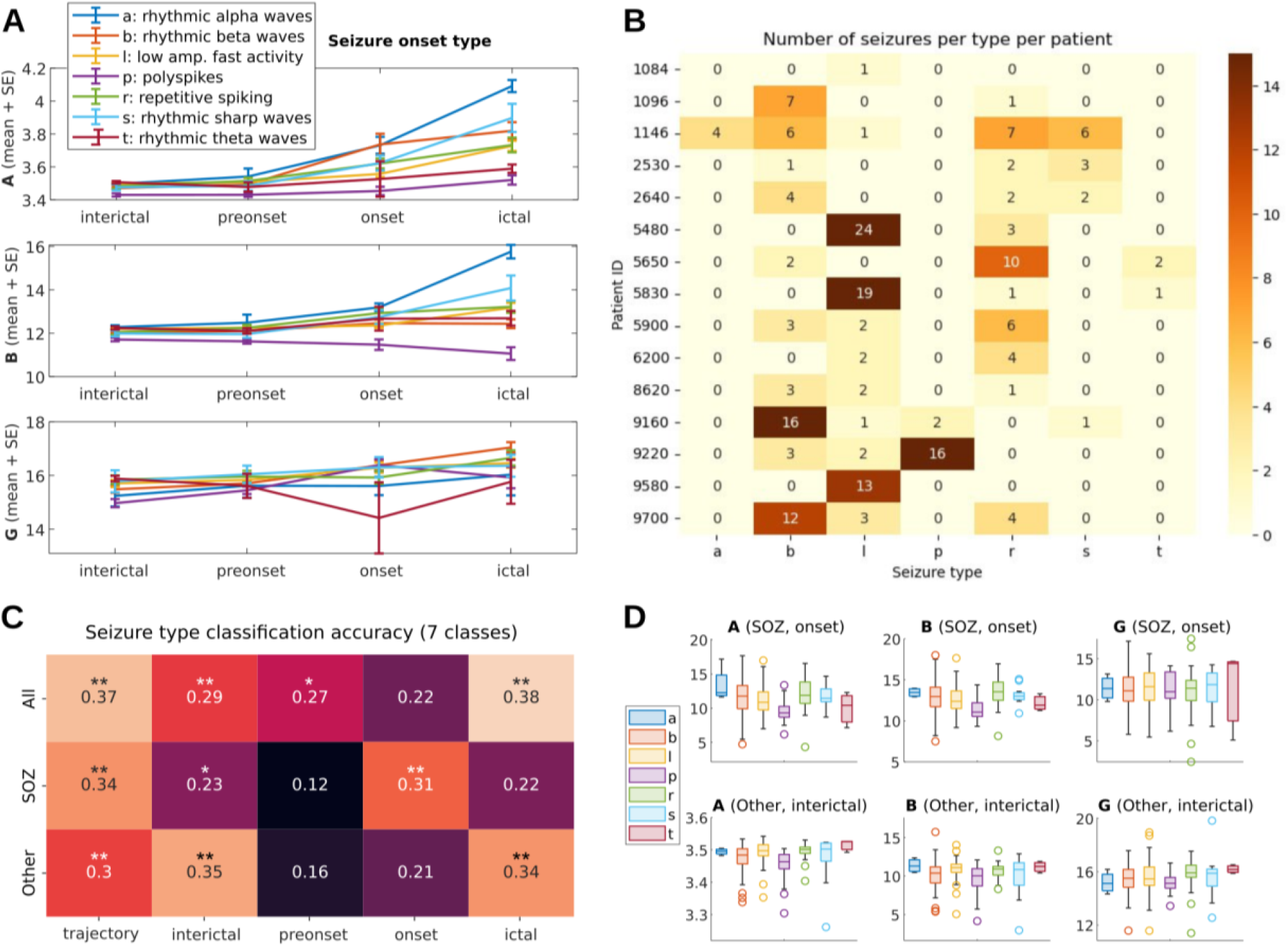
Excitation/inhibition changes across different seizure types. (A) A, B, G evolution averaged per seizure type. Different types of seizures clearly vary in strength and time point of change. (B) Number of seizures per type and patient. Note that there were different amounts of seizures per type and patient, with type “b”, “l”, and “r” being the most common across patients. (C) Classification performance across brain areas (All, i.e. including both SOZ and Other channels; SOZ; Other) and time points (trajectory, i.e. including all 4 time points; interictal; preonset; onset; ictal). Best classification performance was achieved by (All /trajectory) and (All / ictal). SOZ channels achieved better classification performance than All or Other channels in the onset time window, however not during seizure (ictal). Interestingly, both SOZ and Other channels classified seizure types already in the interictal time window. (D) Model fitting results of excitation (A) and inhibition (B and G) per seizure type, for SOZ channels during onset (stronger differentiation expected) and Other channels during interictal (weaker differentiation expected).

Importantly, there were different numbers of seizures per type and patient (Fig. 2B). This led us to use a balanced accuracy measure, equally considering the types, and to use the leave-one-seizure-out cross-validation approach instead of a leave-one-patient-out approach, in which some seizure types, e.g. type “a”, “p” and “t” present in only one or two patients would not have had enough data to train the classifier during leave-one-patient-out.

We found that the classifier could identify the different types of seizures based on their A, B, G trajectory (Fig. 3A, all time points) with a balanced accuracy of 0.37, which was significantly above chance in the leave-one-out cross-validation at the seizure level, (*p*_*FDR*_ = .004; see Fig 3C: All, trajectory). Note that the chance level in a 7-way classification task is 100 / 7 = 14.29. A control analysis (see Supplement S3 and S4), where we conducted a leave-one-out cross-validation at the patient level, was not significant with balanced accuracy of 0.14 and *p*_*FDR*_ > .05. Thus, sample size restrictions and patient-specific effects might have biased classification results.

Fig. 3C also shows the classification performance when considering only selected time points (interictal, preonset, onset, ictal) or brain locations (SOZ, Other). Best classification performance was achieved using all time points, or ictal time window only, in All channels. This implies that the synaptic brain mechanisms determining different seizure onset types extend beyond SOZ channels and the onset time window. However, in the onset time window in particular, only SOZ channels achieved significant classification performance while All and Other channels did not (Fig. 3C: onset). During seizure (Fig. 3C: ictal), All and Other channels achieved significant classification while SOZ channels did not (anymore). Taking into consideration that some of Other channels will show seizure activity later, one possible interpretation is that seizure onset mechanisms in channels outside SOZ are similarly distinct as those inside, just time delayed. That the seizure onset patterns in SOZ channels do not (anymore) differ at seizure time point implies that seizure mechanisms are unspecific to seizure onset types. Importantly, Other (as well as SOZ) channels could distinguish the different types of seizure not only during seizure, but already in the interictal time window. This suggests that the type of seizure is determined already at least 30 seconds before the occurrence of a seizure, and not only in the SOZ, but also in channels outside.

We then studied classification performance separately for A, B, and G parameters, to study which of the neuronal populations involved (A: excitatory pyramidal cells, B: peri-dendritic slow inhibitory interneurons, and G: peri-somatic fast inhibitory interneurons) would be most discriminative between the different seizure types. We found that B was the strongest discriminator among the different types of seizure onsets, across time points and brain areas, with a maximum balanced accuracy of 0.31 (*p*_*FDR*_ = .004; see Supplement S2).

To exemplify the mechanistic insight gained by the modeling procedure, we studied two of the most common seizure onset types in detail. Fig. 4A shows examples of these two types: the low-frequency fast activity (“l”) and repetitive spiking (“r”) onsets. For both types, variance in excitation/inhibition levels increased from interictal to ictal, with A, B, G values being confined in a small area of the A, B, G parameter space (Fig. 4B).

**Figure 4:**
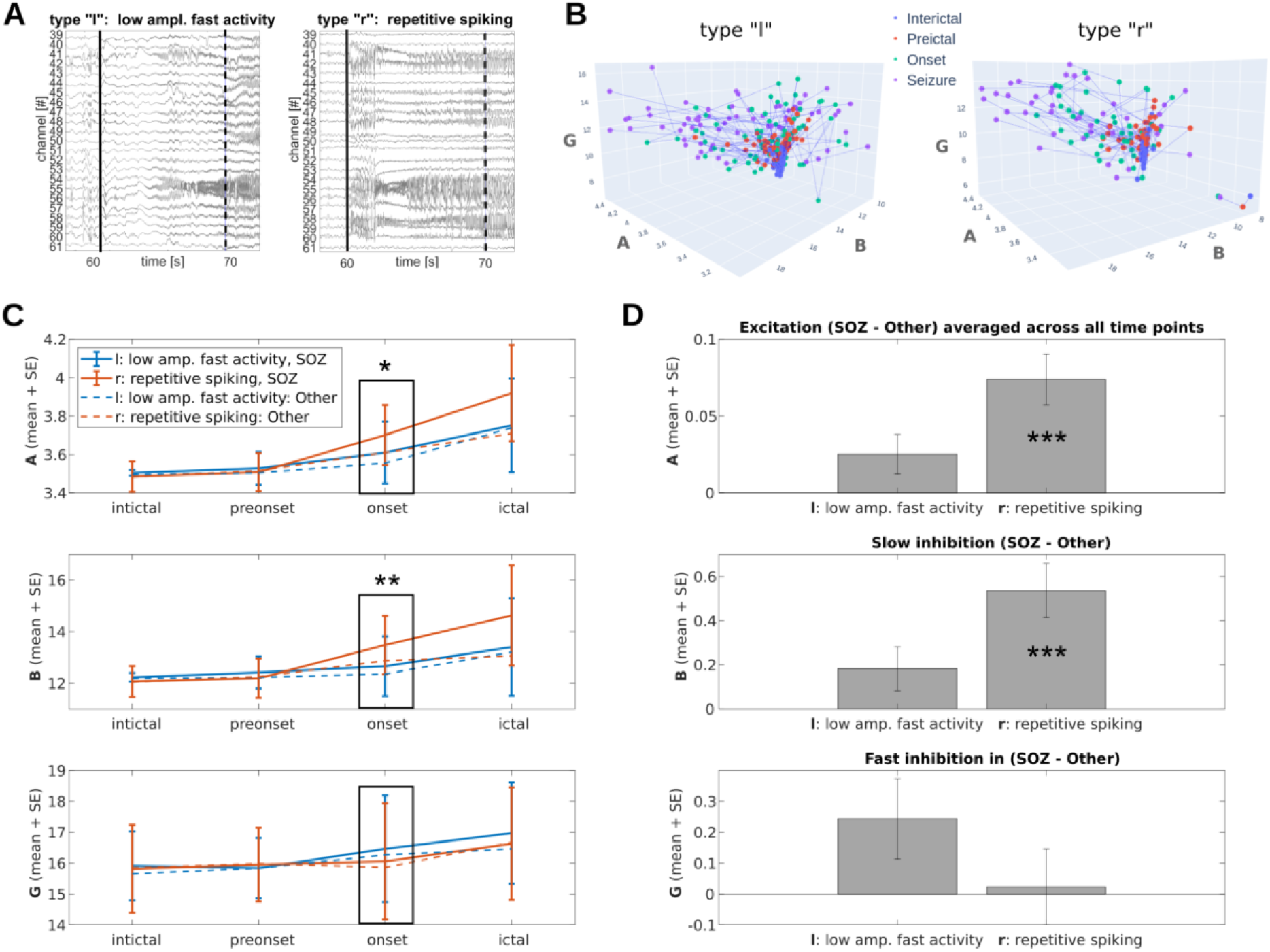
Brain mechanisms involved in “l” and “r” type seizures. (A) Example seizure recording of type “l”: low amplitude fast activity and type “r”: repetitive spiking. The vertical line marks clinically marked seizure onset, the dashed line marks the end of the onset interval as considered in this study (10 seconds). While type “l” shows high frequency activity and an initially small amplitude, type “r” starts with sudden high amplitude spiking, previously described as “low-voltage fast activity onset” and “hypersynchronous onset”, respectively, in the literature. (B) Model fitting results for all seizures of type “l” and type “r”. Seizure transition starts from a confined area in A, B, G space (interictal) and spreads out during seizure transition. (C) A, B, G trajectories averaged across seizures for “l” type (blue) and “r” type (red), in SOZ (solid line) vs. Other (dashed lines). “r” type seizures are associated with an increase of excitation A and slow inhibition B. “l” type seizures only descriptively show an increase in fast inhibition G. (D) Difference between SOZ and Other channels averaged across the four time points (interictal, preonset, onset, ictal), showing SOZ-localization of “r” type seizures (A, B), but not in “l” type seizures. Excitation/inhibition changes in SOZ were generally stronger (SOZ-other > 0).

In line with our hypotheses from the literature, “r” type seizures exhibited stronger excitation A during seizure onset than “l” type seizures (A_r_ > A_l_, *p*_*FDR*_ = .018), which, in turn, at least descriptively showed stronger fast inhibition G (G_r_ < G_l_, *p*_*FDR*_ > 0.05, Fig. 4C). As expected, excitation changes during “l” type seizure transitions were stronger in SOZ than in Other (A_r_ (SOZ - Other) > 0 with *p*_*FDR*_ = .001), while not significant for “r” type seizures (G_l_ (SOZ - Other) > 0 with *p*_*FDR*_ > 0.05 Fig. 4D). Surprisingly however, slow inhibition parameter B was higher during “r” onset (B_r_ > B_l_, *p*_*FDR*_ = .008) and localized in SOZ (B_r_ (SOZ - Other) > 0 with *p*_*FDR*_ = .001).

Finally, in an exploratory analysis, we study a potential correlation of the identified differences in A, B, G evolution per patient (Fig. 1D) and the individual patient characteristics (see Table 1). Unfortunately, the small sample size did not allow a statistical evaluation. Exploratory descriptive results suggest weaker excitation and fast inhibition changes from interictal to ictal in patients with Hippocampal Sclerosis (n = 4) as opposed to without (Fig. 5A), a stronger increase in slow inhibition (B) in patients with an epilepsy focus in the right brain hemisphere (n = 4; Fig. 5B), and overall stronger excitation/inhibition increases in patients that acquired complete seizure freedom after brain surgery (surgery outcome Ia; n = 9) than those who did not (surgery outcome IIb; n = 3; Fig. 5C). However, bigger patient samples are needed to study these potential associations.

**Figure 5:**
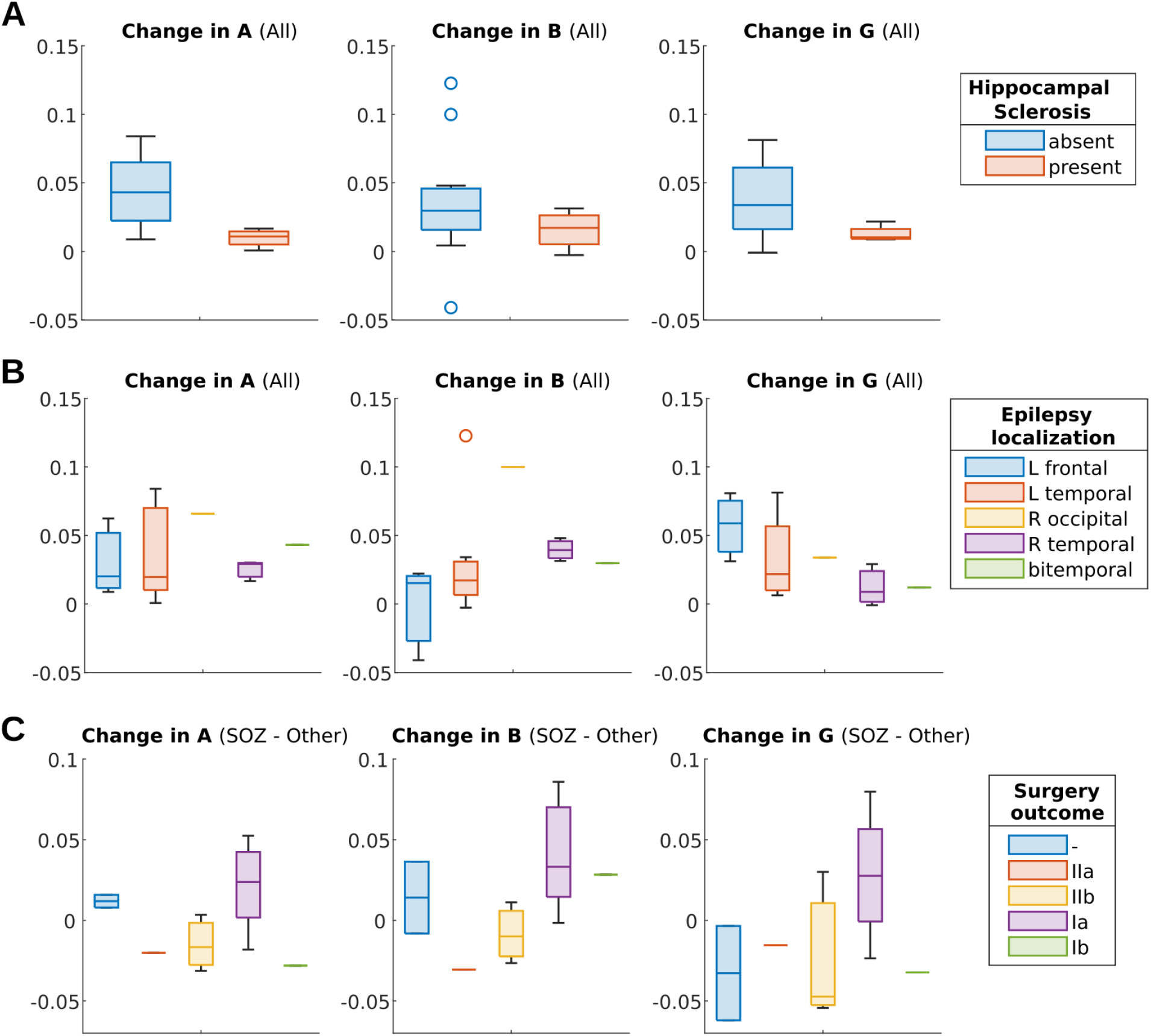
Patient variables and model-predicted excitation/inhibition changes during seizure transition. (A) Averaged excitation/inhibition (A, B and G) changes across all channels and seizures (ictal - interictal) across patients with and without Hippocampal Sclerosis. Patients with Hippocampal Sclerosis (n = 4, red) descriptively show weaker excitation/inhibition changes from interictal to ictal. (B) Same as in A, across patients with different epilepsy localization. Stronger increase in slow inhibition (parameter B) is observed in patients with epilepsy focus in the right brain hemisphere (n = 4). (C) Comparison of respective excitation/inhibition changes between SOZ and Other, calculated as (SOZ - Other), across patients for different surgery outcomes. Patients who acquired complete seizure freedom after brain surgery (surgery outcome Ia; n = 9) descriptively show stronger excitation/inhibition increases than patients with undesirable surgery outcome (IIb, n = 4). Importantly, however, the patient sample was too small for meaningful statistical testing.

## DISCUSSION

This study’s results demonstrate that the predicted synaptic inhibitory and excitatory gain changes during 205 seizure transitions in iEEG significantly differ between seven clinically marked seizure onset patterns.

While across the patterns both excitation and inhibition clearly increased during seizure transition, seizure types differed in magnitude of change and neuronal population involved (excitatory pyramidal cells, peri-dendritic slow inhibitory interneurons, and peri-somatic fast inhibitory interneurons), with B parameter showing strongest discrimination power (however best classification achieved by combining A, B, and G). Interestingly, the types of onsets could be distinguished not only in clinically marked SOZ but also in non-onset channels, and even 60 – 30 seconds before seizure onset.

Our findings support and extend previous literature on the distinct mechanisms underlying different seizure onsets. Because the types of previously described seizure onset patterns and the terminology used to describe them is vast and variable (Abdallah et al., 2024), previous studies on mechanisms often focused on the distinction of the two most commonly observed seizure onset patterns: low-voltage fast (LAF) and high-amplitude slow (HAS; corresponding to low-voltage fast activity and hypersynchronous onset in the introduction), studying their associated initiation dynamics, spatial recruitment, and tissue excitability (Amiri et al., 2019; Enatsu et al., 2012; Perucca et al., 2014; Wang et al., 2017). In terms of correspondence with the seven clinically labeled seizure types in our study, LAF type corresponds to “l: low amplitude fast activity” and HAS type corresponds most closely to “r: repetitive spiking”. Also in our study, type “l” and “r” (next to “b: rhythmic beta waves”) were the most common types of seizures.

Previous studies found that LAF seizures start from small, isolated patches of localized activity, initially controlled by surround inhibition, that gradually expand and merge over time (Wang et al., 2017). This activity reflects the presence of microdomains with localized oscillations, which slowly coalesce and invade surrounding tissue. The cortical tissue surrounding LAF seizure onset exhibits relatively low excitability, which may constrain rapid seizure propagation (Schevon et al., 2012; Wang et al., 2017). Modeling studies have further highlighted the importance of inhibitory gain and synaptic dynamics in shaping LAF seizure onset, particularly pointing to altered inhibitory transmission as a key contributor to seizure generation (Wendling et al., 2002, 2005).

In contrast, HAS onsets indicate a global, systemic shift into the seizure state, initiated by a focal triggering event (Wang et al., 2017). This transition is enabled by a higher excitability of the surrounding cortex, which promotes rapid and widespread synchronization. In line with that, HAS onset seizures have been linked with higher cortical excitability within the seizure onset zone than LAF seizures (Enatsu et al., 2012)

For our study, the two hypotheses of seizure generation in LAF and HAS predicted a stronger excitation A and a potential drop in inhibition parameters G and B, reflecting global hyperexcitability in “r” type HAS seizures. For “l” type LAF seizures, on the other hand, excitation would only moderately increase with a preserved inhibition, where fast inhibition G supports the fast oscillatory activity.

We indeed found that “r” type seizures had higher excitation and lower fast inhibition in the onset interval than “l” type seizures (see Fig. 37C). Furthermore, excitation/inhibition changes from interictal to ictal in “r” type seizures seemed to be more SOZ-localized, than “l” type seizures (see Fig. 38D). Interestingly, however, slow inhibition parameter B, which emerged to be the strongest discriminator between different seizure types, acted against our expectations. Instead of low slow inhibition during “r” and high inhibition during “l” type seizures, we found the opposite pattern, with B parameter seemingly following excitation dynamics in parameter A (see Fig. 38D). This finding challenges the simple view that HAS seizures result uniformly from a loss of inhibition. In line with this, Cossart et al. (2001) demonstrated in experimental temporal lobe epilepsy that inhibition changes are highly compartment-specific: somatic inhibition was increased while dendritic inhibition decreased, with enhanced somatic inhibition resulting from hyperactivity of somatic-projecting interneurons. Wendling et al. (2002) proposed that depression of somatic inhibition by dendritic inhibitory postsynaptic currents could explain transitions to fast onset activity, suggesting that elevated slow dendritic inhibition might actually suppress the more seizure-protective fast perisomatic inhibition through network-level interactions. In line with that, increasing depolarizing GABA_A_-mediated IPSPs can switch activity from background to fast onset seizure patterns (McArdle et al., 2024). Additionally, under pathological conditions with intense GABAergic activity, GABA_A_ receptor activation can produce paradoxically depolarizing rather than hyperpolarizing effects due to altered chloride homeostasis, which could partially explain the similar behavior of excitation and slow inhibition in this study. Future studies using chloride imaging and cell-type-specific manipulations of dendritic-targeting interneurons could disambiguate whether the elevated B parameter reflects genuinely increased inhibitory conductances with altered polarity, compensatory but ineffective upregulation, or network-level suppression of fast inhibitory circuits.

As potential consequence of the different mechanistic underpinnings, previous studies have found the LAF pattern to be linked to more favorable postsurgical outcomes (Abdallah et al., 2024; Bartolomei et al., 2017; Lévesque et al., 2012; Perucca et al., 2014), even though the literature is not entirely consistent (Wang et al., 2017). A current meta-analysis further questioned the strength of this association, naming size of SOZ and completeness of presumed epileptogenic zone resection as the main predictors of surgical outcome (Lagarde et al., 2016, 2019). We argue that these factors are closely linked to the mechanistic underpinnings of the observed patterns, determining together how easily localized and completely resected the EZ can be, and thus determining surgery outcome. For example, the mechanistic hypothesis of independent patches of localized activity controlled by surround inhibition in LAF aligns with the finding of a larger SOZ, suggesting more widespread initial involvement during seizure generation (Perucca et al., 2014; Velascol et al., 2000). While a larger SOZ might be harder to resect completely, its patched, localized nature makes it easier to determine the onset areas. On the other hand, the truly focal instead of regional onset topography in HAS could benefit complete resection; yet, SOZ localization might be trickier, as the hypersynchronous activity quickly spreads to other areas. Given the stronger support for better surgery outcomes in LAF as compared to HAS, we speculate that the localization of SOZ areas, which is easier in LAF, might be a bigger bottleneck and thus more crucial to the prediction of surgery outcome than the size of the SOZ. Lobe-specific localization, as in medial temporal lobe epilepsy, might further be an important influencing factor. HAS seizures in mesial temporal lobe epilepsy have been associated with greater neuronal degeneration and delayed propagation to contralateral regions (Velascol et al., 2000). The more localized pathology (e.g., Hippocampal Sclerosis) and slower propagation might help SOZ delineation.

In line with the literature, patients with predominant “l”/LAF type seizures (n=4) in our study had larger SOZ (mean ± STD number of channels: 15.8 ± 6.4) than patients with “r”/HAS type (n=3; mean ± STD number of channels: 9.3 ± 6.7). In the LAF group, 3 out of 4 patients achieved seizure freedom after surgery, and in the HAS group, 2 out of 3 did, so in both groups, the surgery outcome was similar. However, there was a tendency for more SOZ-localized excitation/inhibition changes in patients with good surgery outcomes, and a stronger excitation/inhibition modulation in general in patients without Hippocampal Sclerosis as opposed to with. Note, that the small sample size did not allow for a thorough investigation of patient sub-groups. We therefore encourage future studies in bigger sample sizes.

Importantly, model-identified excitation/inhibition parameters differed among the types of seizures already during the interictal interval, appealing to a potential usefulness of model-driven seizure onset type labeling, but also implying the pre-determined nature of specific seizure onsets. As seizure types are variable within patients, the dynamics that lead to one onset pattern or another could be governed by a brain dynamic on a slower time scale, predisposing the network to one seizure type or another. To study this slow dynamic, it could, for example, be interesting to investigate whether seizures that occur closer together in time are more likely to share the same seizure onset pattern.

In conclusion, this study showed how different seizure onset patterns in focal epilepsy reflect underlying differences in synaptic communication, regulating the excitability of neuronal populations and spatial organization. Understanding these patterns not only advances our theoretical knowledge of seizure generation but also carries direct implications for the localization of the epileptogenic zone and the prediction of surgical success. Future studies could further study the impact of the mechanistic underpinnings of seizures on other treatment approaches in epilepsy, such as anti-seizure medication and brain stimulation, where the predictions of optimal treatment parameters should also (if not even more so) be directly linked to the underlying mechanisms of seizure generation.

## Data Availability

All data produced are available online at

https://campus-technologies.de/en/the-european-epilepsy-database-2/

## ACKNOWLEDGEMENT

This work was supported by the ERDF-Projects Brain dynamics (No. CZ.02.01.01/00/22_008/0004643, a Lumina-Quaeruntur fellowship (LQ100302301), and the long-term strategic development financing of the Institute of Computer Science (RVO:67985807) of the Czech Academy of Science.

## CONFLICT OF INTEREST

None of the authors has any conflict of interest to disclose.

## Notes

### Competing Interest Statement

The authors have declared no competing interest.

### Author Declarations

The study used (or will use) ONLY openly available human data that were originally located at: https://campus-technologies.de/en/the-european-epilepsy-database-2/. Prior to dissemination through the database, all individual‑level data were pseudonymized in accordance with data‑protection and research‑ethics requirements, such that direct identifiers were removed, and data were managed under a controlled‑access framework.

