## SUPPLEMENT for "Distinct Synaptic Excitation–Inhibition Mechanisms Underlie Clinically Defined Seizure Onset Patterns"

### Model-identified excitation/inhibition changes differentiate seizure onset patterns in intracranial recordings of epilepsy patients

Isa Dallmer-Zerbe (1), Anna Pidnebesna (1,2), Jaroslav Hlinka (1,2)

1 Institute of Computer Science, Czech Academy of Sciences, Prague, Czech Republic
2 National Institute of Mental Health, Klecany, Czech Republic

#### S1: Classification results, main manuscript:

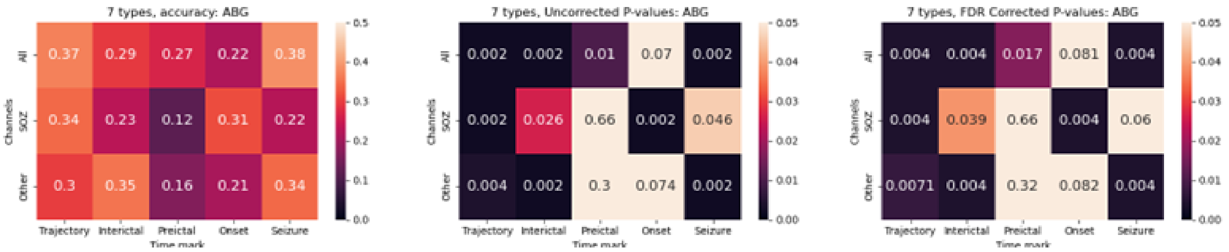


#### S2: Classification results, separately for A, B, and G:

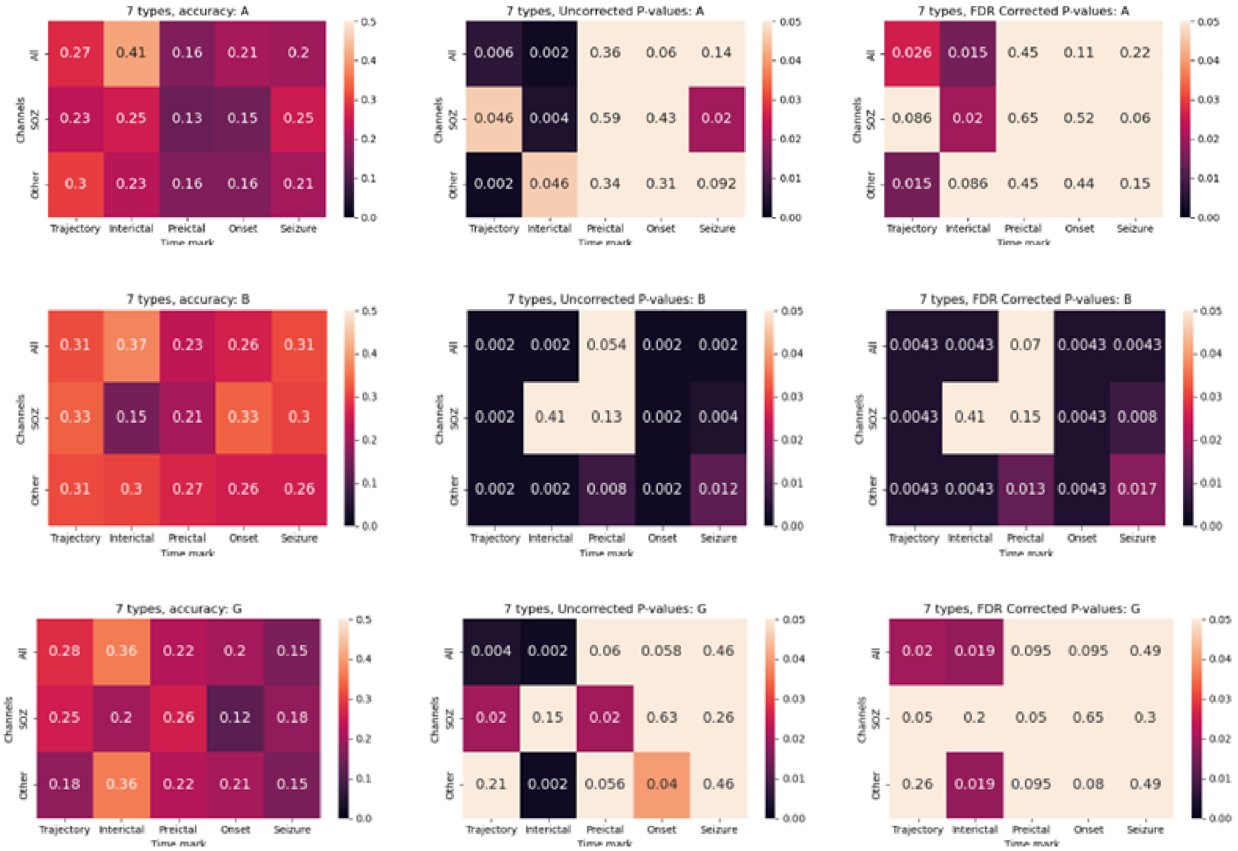

**S3: Classification results for leave-one-patient-out control analysis:**

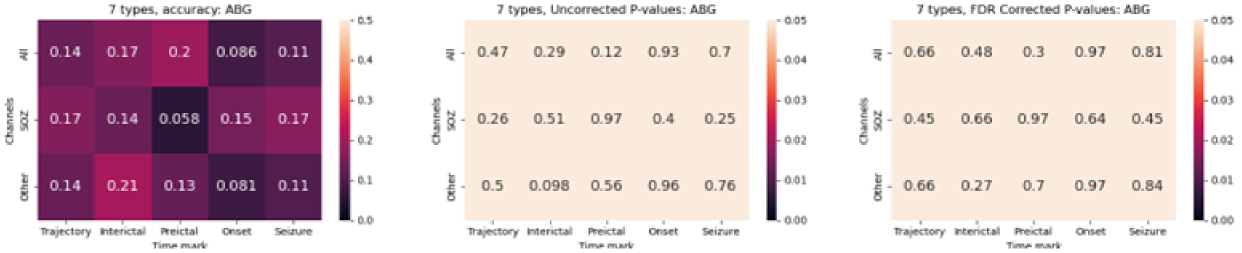


**S4: Classification results for leave-one-patient-out control analysis, separately for A, B, and G:**

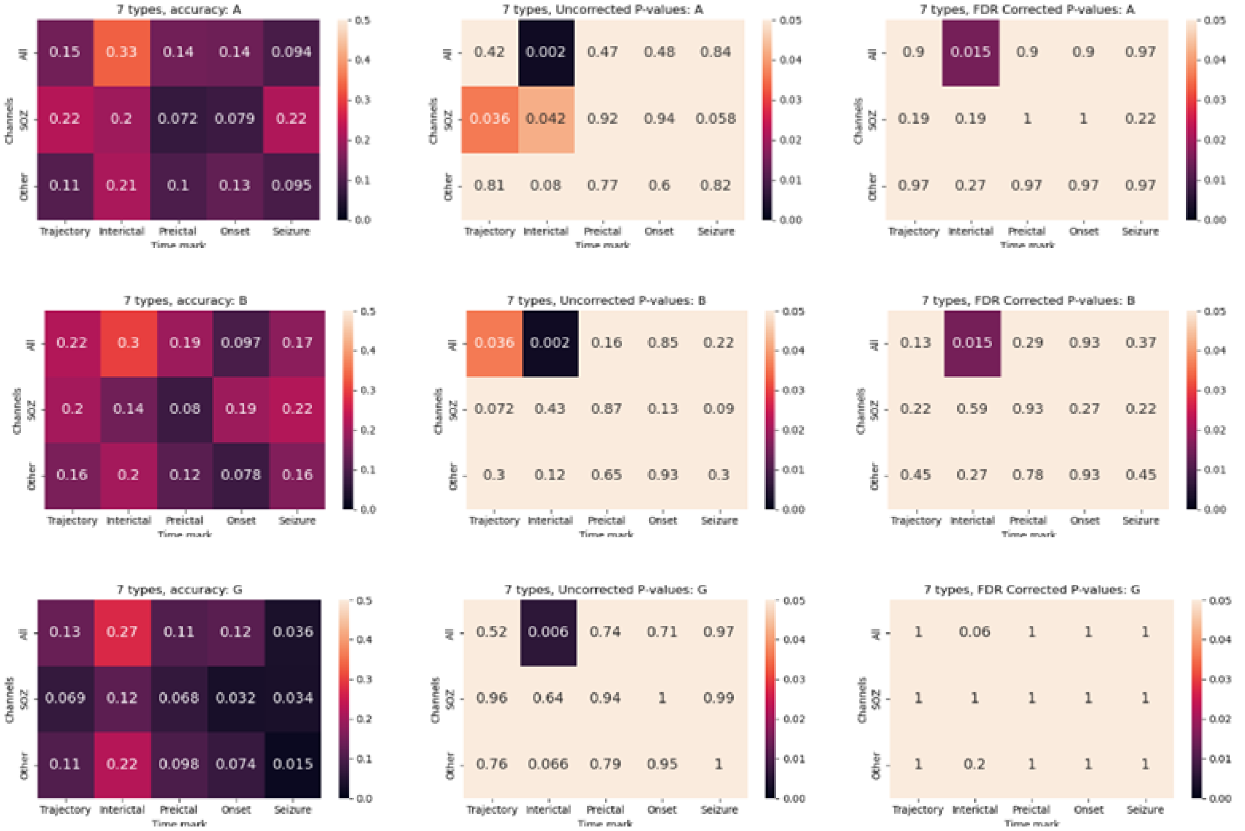
